# Novel Coronavirus in Nigeria: Epidemiological analysis of the first 45 days of the pandemic

**DOI:** 10.1101/2020.04.14.20064949

**Authors:** Oyelola A. Adegboye, Adeshina I. Adekunle, Ezra Gayawan

## Abstract

**Background:** On December 31, 2019, the World Health Organization (WHO) was notified of a novel coronavirus in China that was later named COVID-19. On March 11, 2020, the outbreak of COVID-19 was declared a pandemic. The first instance of the virus in Nigeria was documented on February 27, 2020.

**Methods:** This study provides a preliminary epidemiological analysis of the first 45 days of COVID-19 outbreak in Nigeria quantifying. We estimated the early transmissibility via time-varying reproduction number based on Bayesian method that incorporates uncertainty in the distribution of serial interval (time interval between symptoms onset in an infected individual and the infector) and adjusted for disease importation.

**Findings:** By April 11, 2020, 318 confirmed cases and 10 deaths from COVID-19 have occurred in Nigeria. At day 45, the exponential growth rate was 0.07 (95% Confidence Interval (CI): 0.05 – 0.10) with doubling time of 9.84 days (95% CI: 7.28 – 15.18). Separately for travel related and local cases the doubling time was 12.88 days and 2.86 days, respectively. Furthermore, we estimated the reproduction number for each day of the outbreak using three-weekly window while adjusting for travel related cases. The estimated reproduction number was 4.98 (95% CrI: 2.65 – 8.41) at day 22 (March 19, 2020), peaking at 5.61 (95% CrI: 3.83 –7.88) at day 25 (March 22, 2020). The median reproduction number over the study period was 2.71 and the latest value at April 11, 2020 was 1.42 (95% CI: 1.26 – 1.58).

**Interpretation:** These 45-day estimates suggested that cases of COVID-19 in Nigeria have been remarkably lower than expected and the preparedness to detect needs to be shifted to stop local transmission.

**Funding:** None

## Introduction

The novel severe acute respiratory syndrome coronavirus 2 (SARS-CoV-2), also known as COVID-19, emerged in the city of Wuhan, China in late December 2019 and was declared a global pandemic by the World Health Organization (WHO) on March 11, 2020.^1^ In the time since, the disease has quickly spread to all continents and, to date, over 1.6 million cases have been recorded with a fatality rate of 6.19% noted on April 11, 2020.^2^

Thus far, the risk of COVID-19 importation from Europe to Africa is higher than the risk of importation from China.^3^ In their study, Martinez-Alvarez et al ^4^ compared early transmission of COVID-19 (within 6 days after the first cases were detected) in selected countries and observed a rapid spread of the virus in some West African countries than in Europe. ^4^ The situation could be worse than what is being reported as most African countries are inadequately prepared for disease outbreak due to poor disease surveillance and response systems as well as inadequate and overstretched health facilities and services. However, African countries with the highest importation risk have been found to possess a high capacity to respond to outbreaks.^5^ As of April 11, 2020, a total of 13,814 confirmed cases and 747 deaths from COVID-19 have been documented in Africa.

Although the first case of COVID-19 in Nigeria was detected on February 27, 2020, this did not lead to an immediate outbreak. The epidemic trajectory has been slow, in part, due to the public health interventions implemented in Nigeria which have reduced both local transmission and importation. ^6,7^ A series of immediate interventions were put in place by the government of Nigeria in response to COVID-19. An international travel ban was imposed on 13 countries on March 20, 2020. Additionally, school and university closures were implemented early and restriction on movements within and outside of major cities were enforced.

In this article, a preliminary epidemiological analysis of the first 45 days of the COVID-19 outbreak in Nigeria is provided. With an increase in COVID-19 importation into Nigeria, large disease outbreak is imminent as this is consistent with observed cases in countries that are epicentres. One key variable of measuring transmissibility of infectious diseases is the effective reproduction number (*R*_*eff*_) which is similar to the basic reproduction number (*R*_0_). The basic reproduction number (*R*_0_) is the average number of secondary cases that arises when one primary case is introduced into an uninfected population. ^8^ Travel has remained a major source of concern for the current COVID-19 pandemic; therefore, early transmissibility of COVID-19 diseases is quantified in Nigeria using sequential Bayesian method, adjusted for disease importation.

## Methods

In this study, the daily number of confirmed cases of COVID-19 were collected from publicly available data of the outbreak situation report of the Nigeria Centre for Disease Control (NCDC) ^7^ and the World Health Organization daily situation reports. ^2^

The real time growth of COVID-19 in the first 45 days was estimated by fitting exponential curves to the daily counts and its changes in time based on log-linear Poisson regression model. Transmissibility of the disease measured by the effective reproduction number (*R*_*eff*_) was estimated from the epidemic curve. In order to account for the effect of disease importation (delay window), sequential Bayesian method was used to estimate time-varying *R*_*eff*_ from the incidence series ^9^. Previous studies have estimated the mean serial interval (time interval between symptoms onset in an infected individual and the infector) of COVID-19 to be 7.0 (5.8–8.1) days, with a standard deviation of 4.5 (3.5–5.5) days ^10^. Therefore, a shifted Gamma distribution with a mean of 7.0 days and standard deviation of 4.5 days was assumed, with shift 1 day used for distribution of the serial interval.

A sensitivity analysis was carried out to investigate the effect of changes in the sliding window on cases *R*_*eff*_. All analyses were done in R software version 3.6.2 ^11^ using packages incidence ^12^ and EpiEstim ^9^.

## Results

Between February 27, 2020 and April 11, 2020 (a 45-day period), a total of 318 confirmed cases and ten COVID-19 deaths (case fatality rate of 3.14%) were recorded in Nigeria. Figure 1 presents the cumulative number of confirmed cases over time and the geographical coverage of the disease. The temporal trend of incidence shows an exponential growth. The first case of COVID-19 occurred in Lagos state, the economic hub of the country, which has remained the focus of the epidemic in Nigeria. About 72% of cases have been reported from the duo of Lagos and the Federal Capital territory (FCT). More than 47% of the cases are travel related and the spread of the disease has been concentrated in the southern region of the country and the FCT (Figure 1).

**Figure 1:**
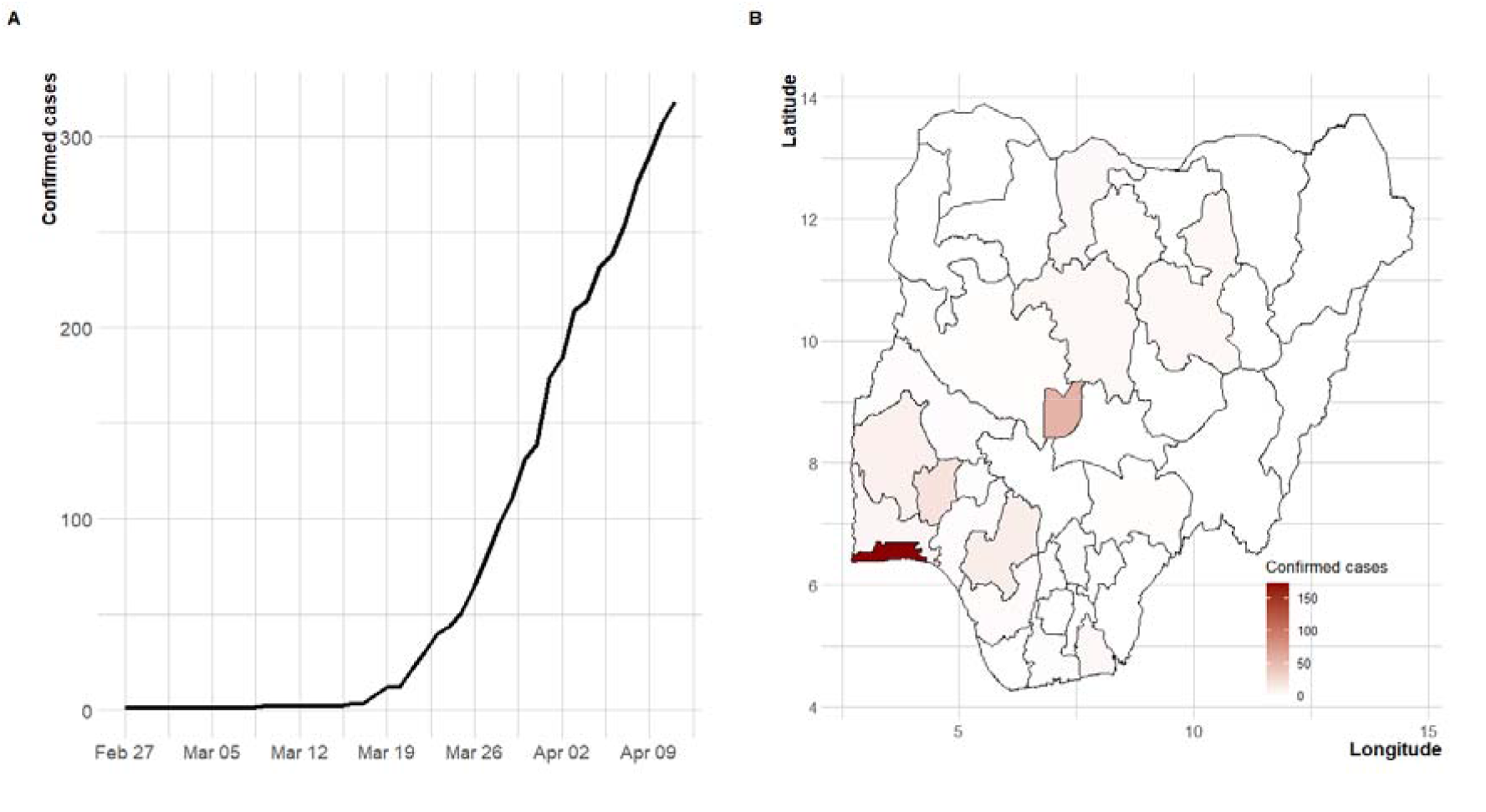
Distribution of COVID-19 disease in Nigeria between February 27, 2020 and April 6, 2020. (A) Time series plot of daily counts, (B) States affected by the disease.

For comparison, the observed daily cases in selected African countries from day 1 up to day 45 are shown in Figure 2 and Figure 3. Initial inspection of the progression indicates that the disease is progressing exponentially more rapidly in some African countries than others (Figure 3). The progression of the disease within the first 45 days is slowest in Nigeria compared with the other African countries. Assessing the case (per 100,000) during the first 45 days (or less in some African countries), the burden of COVID-19 is lowest in Nigeria (0.16 cases per 100,000 as of April 11, 2020) among African countries with the most cases.

**Figure 2:**
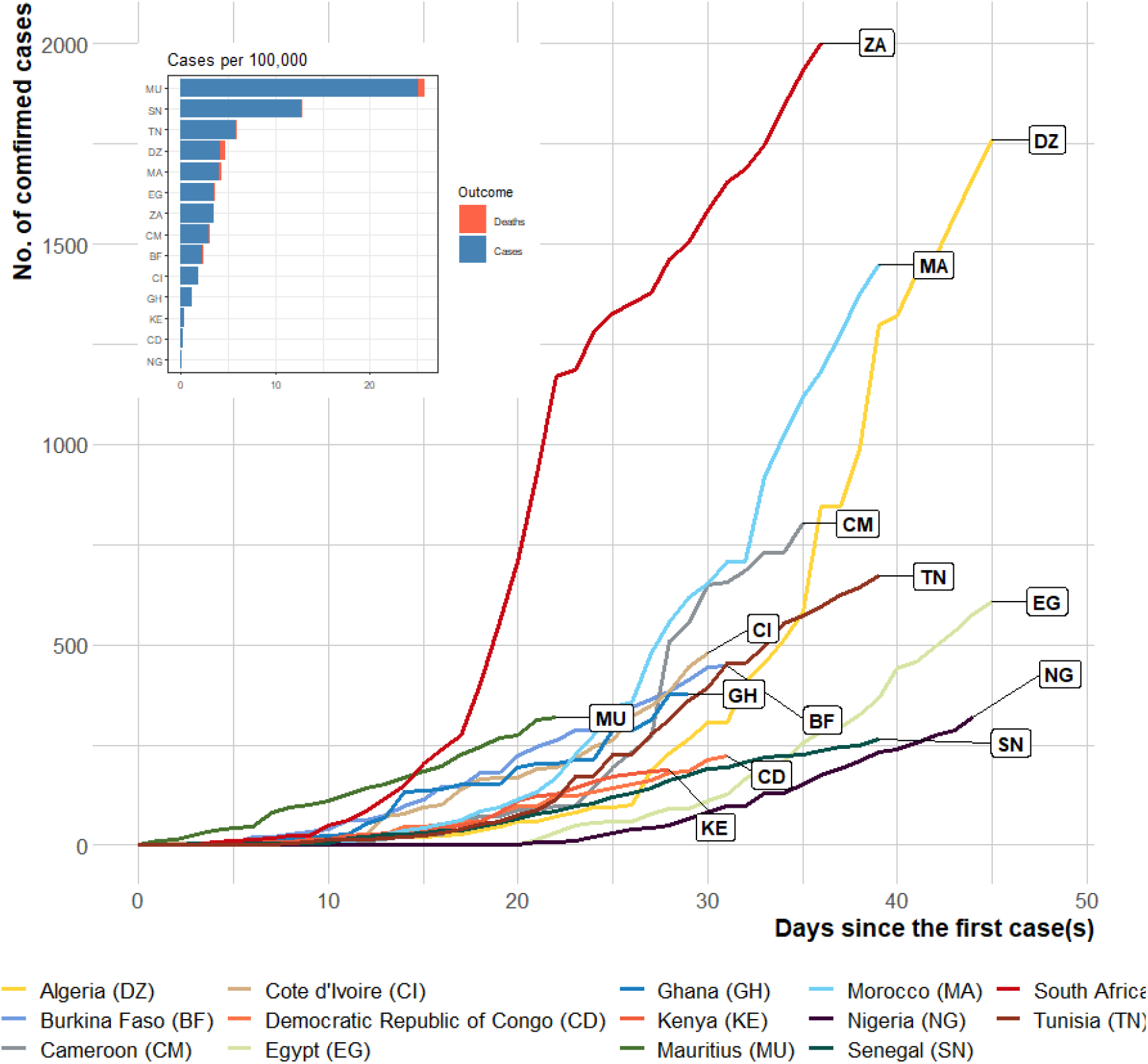
Number of confirmed cases in the first 45 days of COVID-19 importation to selected African countries. Inset: Cases per 100,000 population as at April 11, 2020.

**Figure 3:**
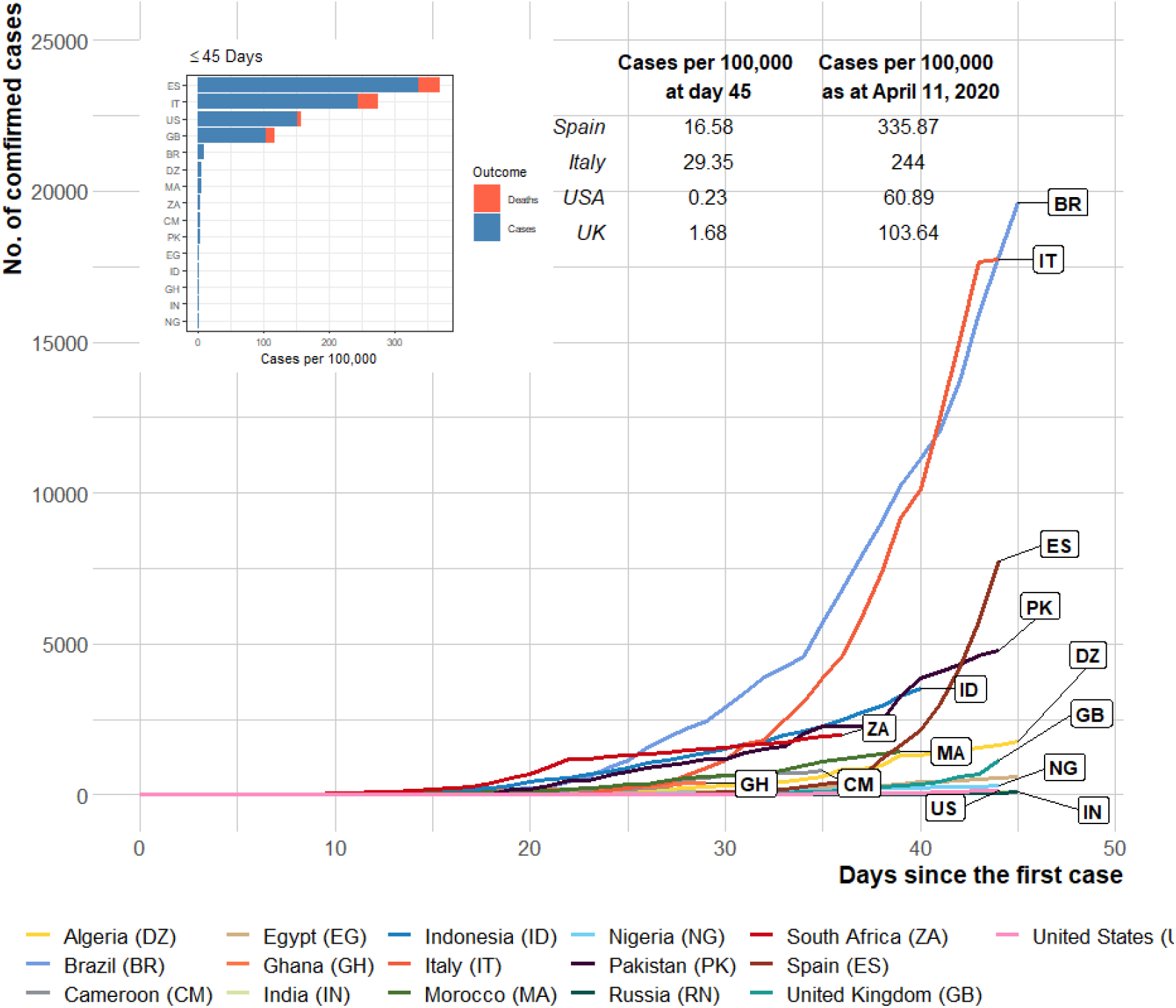
Number of cases in the first 45 days of COVID-19 arrival to selected countries (Inside and outside Africa). Inset: Bar plot showing cases per 100,000 population at day 45. Comparison of cases per 100,000 at day 45 (March 5 for USA, March 15 for UK, March 16 for Spain, March 15 for Italy) and at April 11, 2020 (around a month later).

In comparison with some of the most affected countries outside of Africa, the number of confirmed cases indicates that the disease spread occurred more slowly in Nigeria within the first 45 days (Figure 3). The small number of cases in the US (0.22 cases per 100,000 on March 3, 2020) and high number of cases in Italy (29.35 cases per 100,000 on March 15, 2020) within the first 45 days are notable. However, this value rose to 60.89 and 244.00 cases per 100,000, respectively, as at April 11, 2020 (a month after).

Figure 4 presents the epidemic curves and fitted exponential growth (EG). An EG model was fitted to the overall observed incidence data (Figure 4A) and separately for travel related incidence data and locally transmitted incidence data (Figure 4B). As at April 11, 2020, and at the current growth rate (0.07, 95% CI: 0.05 – 0.10), the doubling time of the epidemic was 9.84 days (95% CI: 7.27 – 15.18), ignoring the travel related cases. On the other hand, when a separate model was fitted to travel related and local incidence data, the doubling time was 24.13 days (95% CI: −18.24 – 7.26) and 5.00 days (95% CI: 0.02 – 30.76), respectively. As shown in Figure 4B, the travel related fitted model is flattening while locally transmission is increasing at a rate of −0.03 (95% CI: −0.10 – 0.04) and 0.13 (95% CI: −0.02 – 0.30), respectively.

**Figure 4:**
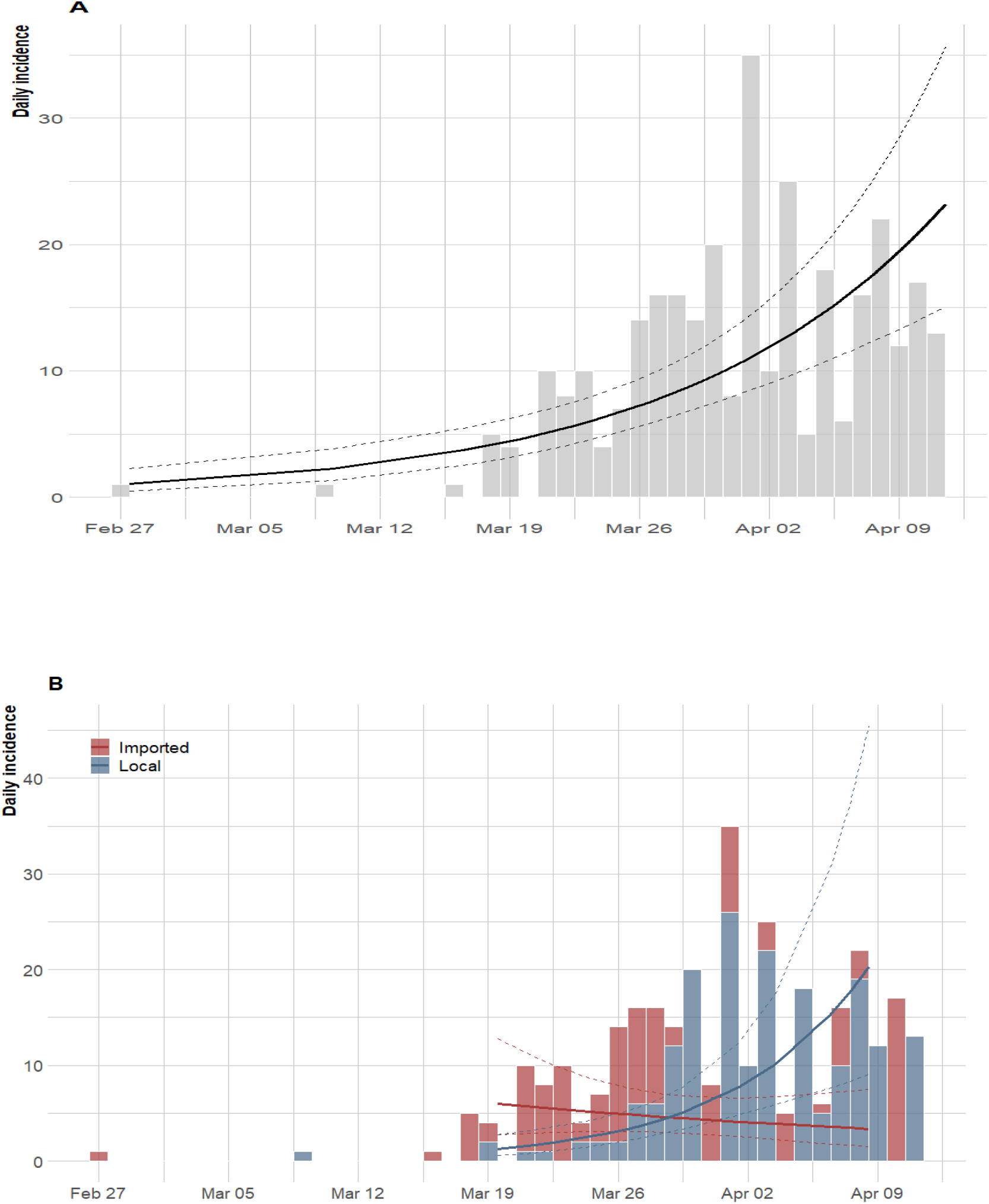
Epidemic curve of the confirmed cases of COVID-19 and the exponential growth fitting. (A) For all daily new cases, (B) disaggregated cases per transmission route as local or imported. The thick line represents the estimate surrounded by the 95% confidence interval (dashed lines).

Furthermore, we estimated the reproduction number for each day of the pandemic using a three-weekly window ending on that day while adjusting for travel related cases. We used three-week sliding window so that the number observed cases of COVID-19 will be at least 12 before starting to estimating the reproduction number. ^9^ Due to imprecise estimates at the beginning of the pandemic, the estimates of time dependent reproduction number are not displayed at the beginning (Figure 5). Following Wu et al ^10^, we assumed that the serial interval and the generation time have the same distribution with mean serial interval of 7.0 (95% CI: 5.8–8.1) days and a standard deviation of 4.5 (95% CI: 3.5–5.5). The time-varying R estimates based on these statistics are presented in Figure 5.

**Figure 5:**
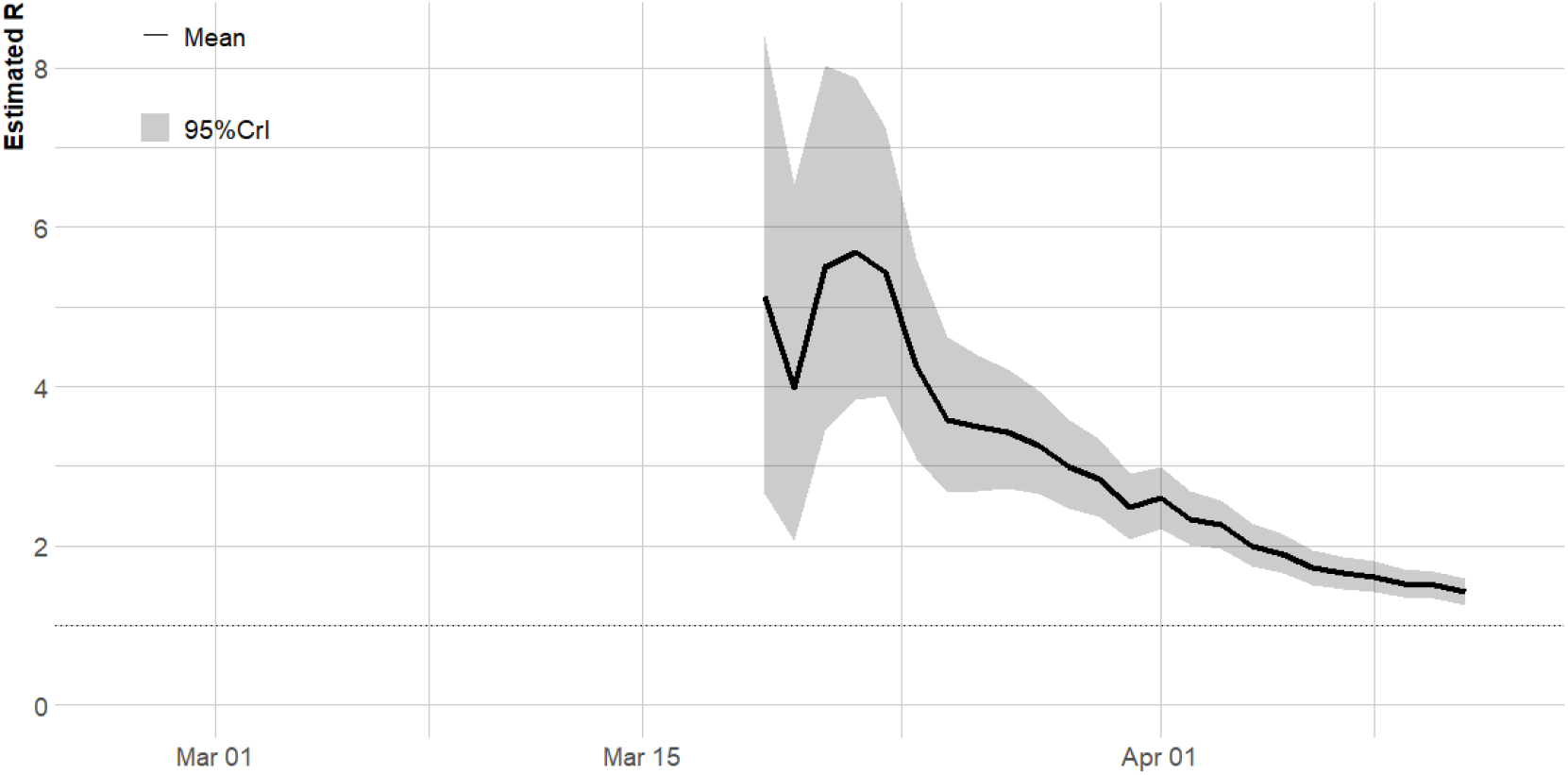
Time-varying reproduction number of COVID-19 in Nigeria based on three-week sliding window ^9,21,22^. That accounted imported and local transmission. The black line represents the posterior median and the grey shaded region represents the 05% credible interval (CL).

The estimates of reproduction number increase rapidly from a median of 4.98 (95% CrI: 2.65 – 8.41) at day 22 (March 19, 2020), reaching a maximum value of 5.61 (95% CrI: 3.83 –7.88) at day 25 (March 22, 2020). The value of R has been decreasing steadily after the country placed an international travel ban on 13 countries on March 20, 2020, it is still above the pandemic threshold of 1. The median R over the study period was 2.71 and the latest value at April 11 was 1.42 (95% CrI: 1.26 – 1.58).

The posterior distribution of SI based on the parametric bootstrap approach with 1000 resamples and 100 simulations is displayed in Figure S4. We assessed that changes to estimating the reproduction number too early in Figure S3. Using a sliding window of one-week with a single case, the latest value at day 45 was 0.99 (95% CrI: 0.81 – 1.19), suggesting that the pandemic is almost over in Nigeria.

## Discussion

This study is the first to provide epidemiological information on the early stages of the COVID-19 outbreak in Nigeria. Although most cases occurred in Lagos state (the economic hub of Nigeria) and the FCT, the disease has spread to 20 states within the first 45 days. We estimated the disease growth rate and the time varying reproduction number for the early cases of COVID-19 in Nigeria. Our modelling results show that COVID-19 is growing and more needs to be done to flatten the curve or squash the spread of the disease. The doubling time for COVID-19 importation and local transmission was 24.13 days and 5 days, respectively. This implies that the epidemic will take a longer time to double based on importation data only but a shorter time for local transmission which calls for additional attention. In particular, the time-varying reproduction number was above one for most of the time period, suggesting that more cases will be recorded in the country in the future.

The first case of COVID-19 was detected in Nigeria on February 27, 2020, but the pandemic trajectory has been slow compared with other countries due to the public health interventions implemented. Among others measures, an international travel ban was imposed on 13 countries on March 20, 2020 followed by a total ban of all international flights in and out of the country, early closure of all schools, universities and worship centres throughout the country and restriction on movements within and outside of major cities which were enforced on March 29, 2020.

As of April 11, 2020, 318 confirmed cases were reported in Nigeria of which about 40% were travel-related ^7^. The case fatality rate was 2.2% and 58 have either recovered or are stable. These figures and the burden of the disease are relatively small when compared with other countries in Africa and Europe (Figure 2). However, considering the geographical landscape of the country, more new cases may be confirmed in the next few weeks.

There are several reasons for late importations of COVID-19 in Africa other than the speculations that COVID-19 may not be viable in temperate regions. One of the reasons is limited international travel.^4^ This may be true as, economically, Nigeria is a developing country and fewer Nigerian tourists and business personnel returning home are expected. Another reason could be a lack of exposure to the virus by Nigerian returnees. When COVID-19 emerged in China, the Chinese government introduced lock downs with people seeking cover and sheltering in different places. Thus, a Nigerian returnee may have limited contacts in China. Hence, the first importation to Nigeria was a resident from Italy rather than China, the original epicentre of the outbreak.

In order to combat emergent infectious diseases in Nigeria, the Nigerian Centre for Disease Control (NCDC) was established in 2011. Its mandate includes detecting, investigating, preventing and controlling diseases of national and international public health importance. The response of the Nigerian government to the 2014 Ebola outbreak was highly commendable and swift. However, the COVID-19 outbreak is quite different from Ebola and may require extra effort to handle the sudden rise in the number of outbreaks within the country. The current outbreak task force is led by the NCDC.

An early comparison with the worse affected European countries within the first six days reveals a rapid acceleration of the pandemic within those countries ^4^, which was not the case for Nigeria and most African countries in the first 30-50 days. Consequently, there is a need for Nigeria to borrow a leaf from these countries and not to relent in the efforts to curb the outbreak. The setting up of nine fully functional COVID-19 laboratories across the country increased the testing capacity to 1500 a day and is a step in the right direction.

Nigeria, being a nation with very peculiar religious tourism and commonplace for large social gatherings such as weddings, needs to enhance “physical” distancing. The spread of COVID-19 has been fuelled by mass migration for religious purposes in some countries. Mass gatherings have been associated with increasing the transmission of virus creating high-risk conditions for the rapid global spread of infectious diseases ^13^. For example, COVID-19 outbreaks were linked to several religious gathering clusters in Singapore ^14,15^, Malaysia ^16^ and South Korea ^17-19^. It is not surprising that for this year, most countries have cancelled religious activities such as the cancellation of Umrah pilgrimage in Saudi Arabia ^20^.

This is an early investigation of COVID-19 cases in Nigeria, as such we acknowledged the following limitations in our study. Firstly, though the data analysed were the official figures released by the Nigeria Centre for Disease Control, the actual cases in the country within the period studied could have been underreported due to low testing capabilities. For instance, as at April 6, 2020, the country was only able to test 5,000 individuals translating to 240 per 100,000 people. Moreover, a lack of proper awareness and fear of stigmatization could have hindered people with suspected cases from coming forward for testing. Secondly, Figure 4B was based on most plausible daily counts from the daily reports therefore, the number of imported cases may not be in real-time, therefore some of patient may have been previously included the cases counted as missing epidemiological information. Lastly,

## Conclusion

This study aims to serve as a reminder to the policy makers, health officers, disease control agencies and the general public that although the number of confirmed cases may be relatively low, the risk is still very high and potentially there could be many asymptomatic cases in the country. Thus far, the intervention in Nigeria has been timely, but the efforts need to be double up. COVID-19 cases in Nigeria are evolving in a similar way to what was observed in the early days of COVID-19 in the USA. With an estimated five hospital beds per 10,000 and four medical doctors per 10,000 population (compared to US with 290 hospital bed and 25.9 medical doctors per 10,000), an outbreak of the magnitude observed in the US will be devastating. The disease is currently concentrated in Southern and North-Central Nigeria, and with ongoing Boko Haram insurgency activities in the North-East of Nigeria, early detection and control of disease outbreaks in the North-East would be very difficult. Public gatherings/events such religious and ceremonial gatherings need to be minimized, and restrictions on movement for an extended period.

## Data Availability

Publicly available data

## Appendix

**Figure S1:**
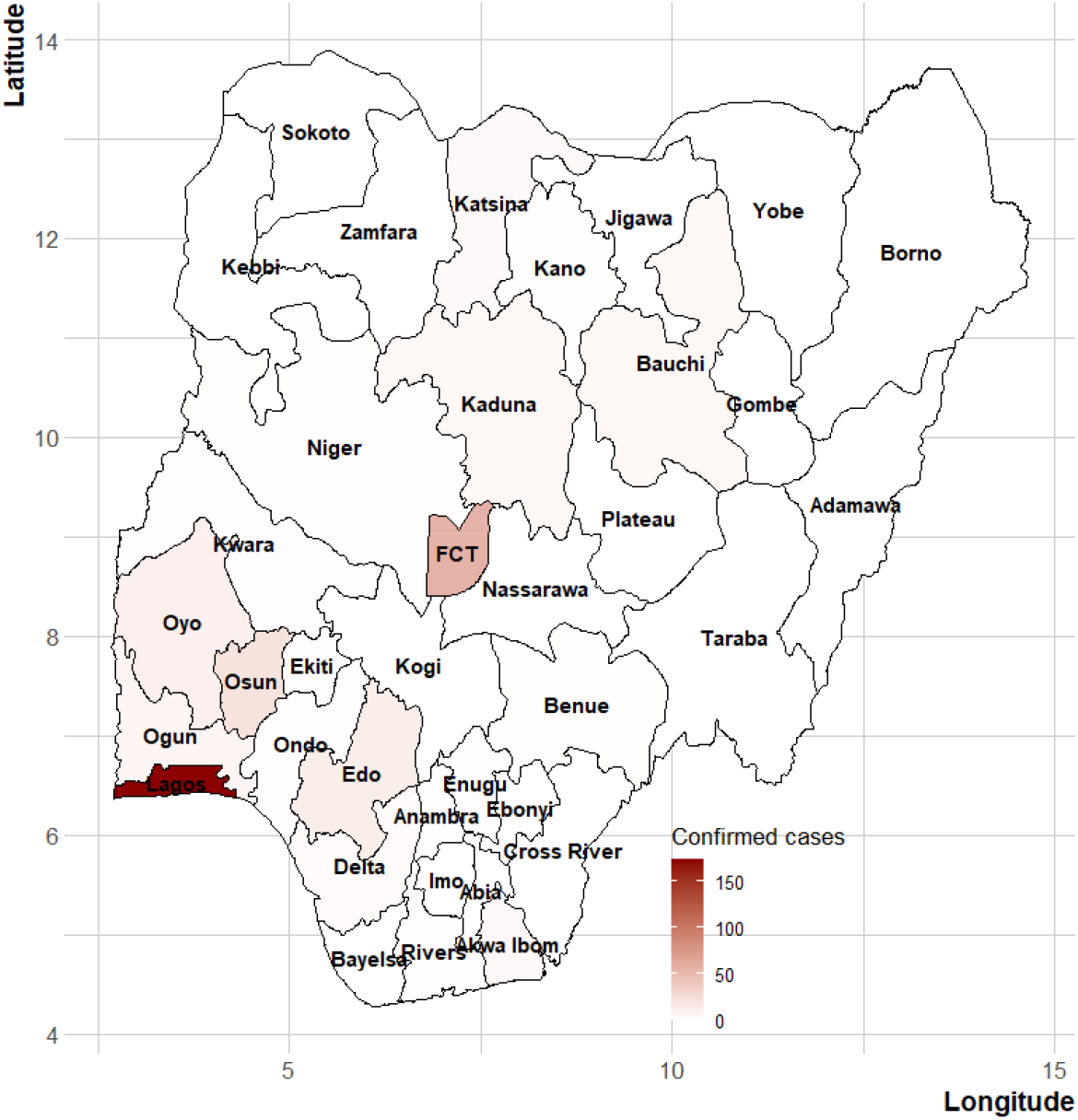
Map of Nigeria showing the distribution of cases of COVID-19 as at April 11, 2020.

**Figure S2.**
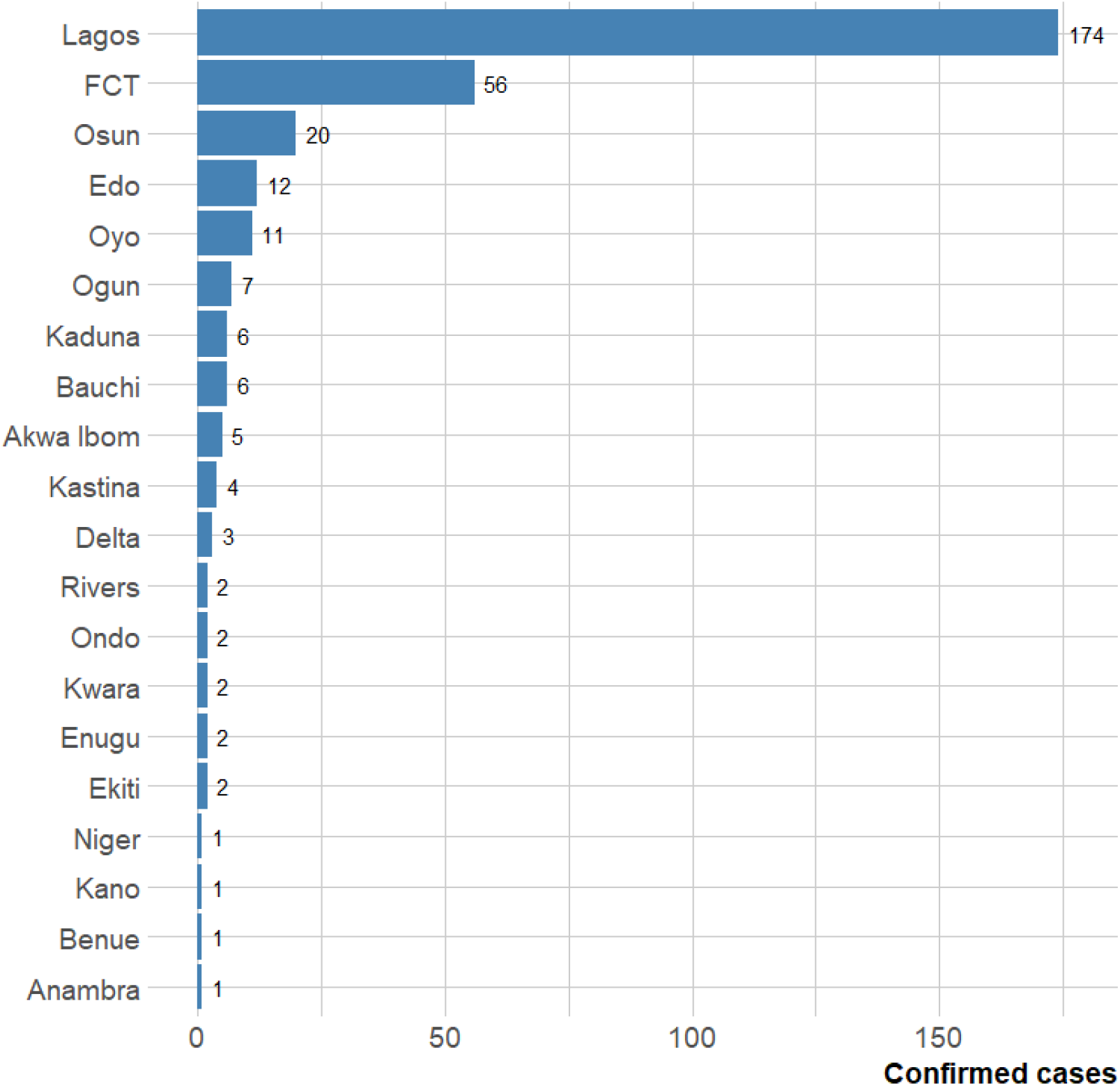
Number of confirmed cases per states in Nigeria.

**Figure S3.**
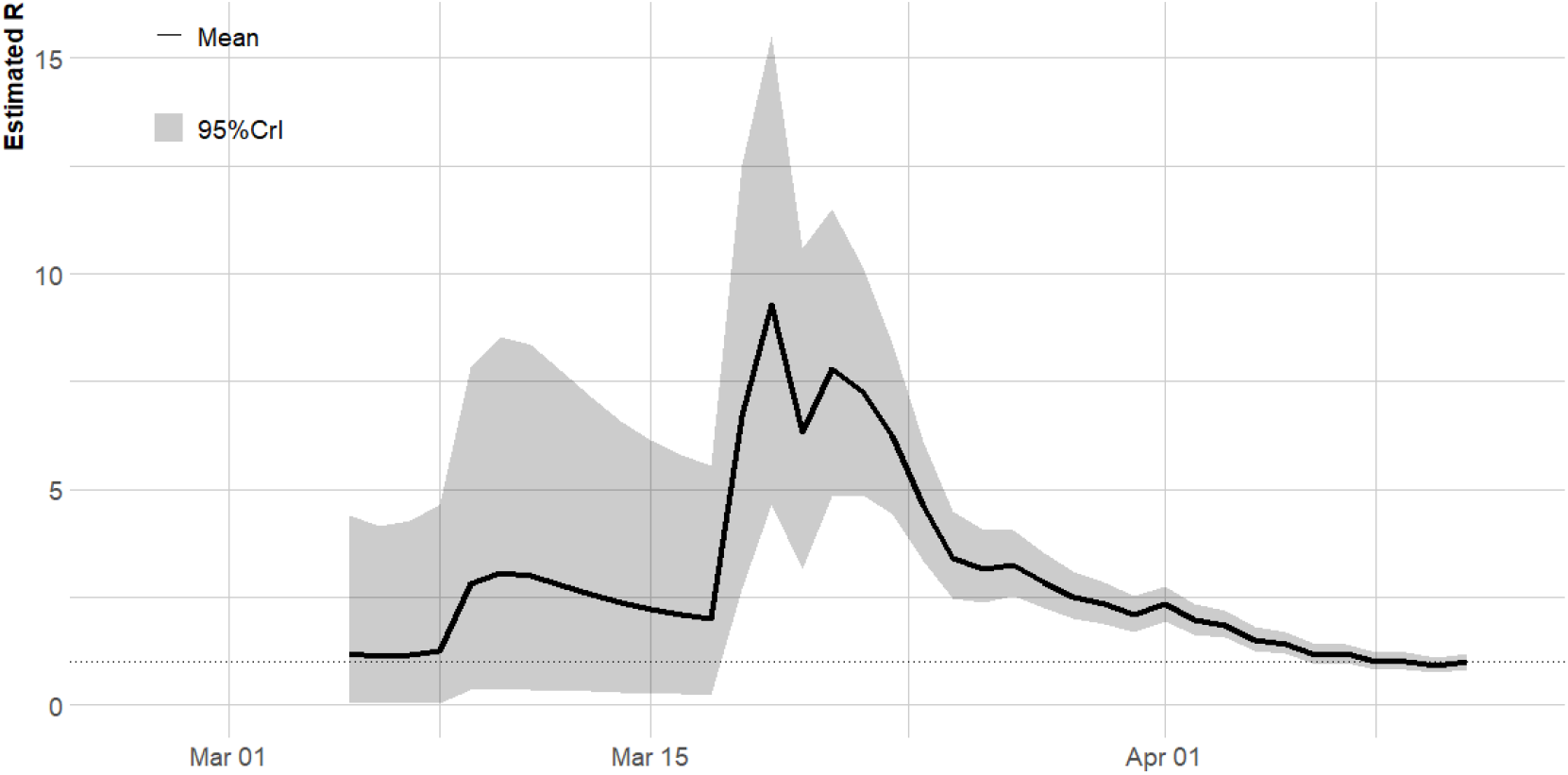
Time-varying reproduction number using one-week window.

**Figure S4.**
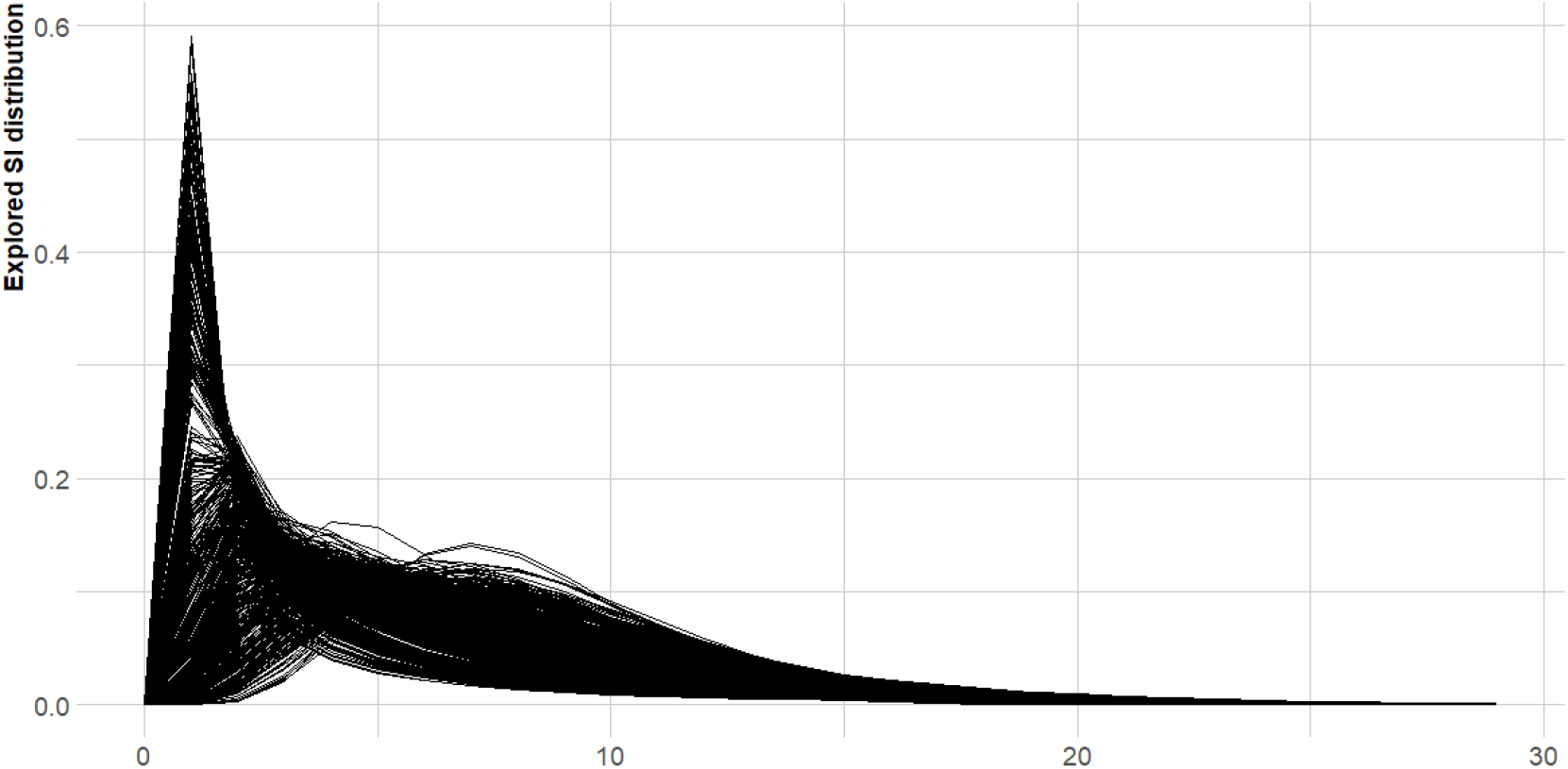
Posterior sample for serial interval distribution for time-varying reproduction number in Figure 5.

## Notes

### Competing Interest Statement

The authors have declared no competing interest.

